# Noninvasive Staging of Lymph Node Status (NILS) in Breast Cancer using Machine Learning: External Validation and Further Model Development

**DOI:** 10.1101/2023.02.11.23285691

**Authors:** M. Hjärtström, L. Dihge, P. O. Bendahl, I. Skarping, J. Ellbrant, M. Ohlsson, L. Rydén

**Author notes:** Shared last authorship (M. Ohlsson, L. Rydén).

## Abstract

**Background:** Most patients diagnosed with breast cancer present with node-negative disease. Sentinel lymph node biopsy (SLNB) is routinely used to stage the axilla, leaving patients with healthy axillary lymph nodes without therapeutic effects but at risk of morbidities from the intervention. Numerous studies have developed nodal (N) status prediction models for noninvasive axillary staging using postoperative data or imaging features that are not part of the diagnostic workup. Lymphovascular invasion (LVI) is a top-ranked predictor of N metastasis; however, it is challenging to assess preoperatively.

**Objective:** To externally validate a multilayer perceptron (MLP) model for noninvasive lymph node staging (NILS) in a large population-based register cohort (n=18 633) while developing a new N MLP in the same cohort. Data were extracted from the Swedish National Quality Register for Breast Cancer (NKBC), 2014–2017, comprising only routinely and preoperatively available documented clinicopathological variables. Furthermore, we aimed to develop and validate an LVI MLP to predict missing values of LVI status to increase the preoperative feasibility of the original NILS model.

**Methods:** Three non-overlapping cohorts were used for model development and validation. Four N MLPs and one LVI MLP were developed using 11–12 routinely available predictors.

Three N models were used to account for the different availabilities of LVI status in the cohorts and external validation in NKBC. The fourth N model was developed for 80% of NKBC cases (n=14 906) and validated in the remaining 20% (n=3727). Three alternatives for imputing missing values of the LVI status were compared using the LVI model. The discriminatory capacity was evaluated by validation area under the receiver operating characteristics curve (AUC) in three of the N models. The clinical feasibility of the models was evaluated using calibration and decision curve analyses.

**Results:** External validation of the original NILS model was performed in NKBC (AUC 0.699, 95% CI: 0.690-0.708) with good calibration and the potential of sparing 16% of patients with node-negative disease from SLNB. The LVI model was externally validated (AUC 0.747, 95% CI: 0.694-0.799) with good calibration but did not improve the discriminatory performance of the N models. The new N model was developed in NKBC without information on LVI (AUC 0.709, 95% CI: 0.688-0.729) with excellent calibration in the hold-out internal validation cohort, resulting in the potential omission of 24% of patients from unnecessary SLNB.

**Conclusions:** The NILS model was externally validated in NKBC, where the imputation of LVI status did not improve the model’s discriminatory performance. A new N model demonstrated the feasibility of using register data comprising only the variables available in the preoperative setting for NILS using machine learning. Future steps include ongoing preoperative validation of the NILS model and extending the model with, for example, mammography images.

**Trial Registration:** Registered in the ISRCTN registry with study ID ISRCTN14341750. Date of registration 23/11/2018.

## INTRODUCTION

Breast cancer is the most frequently diagnosed cancer worldwide; however, its overall prognosis is good [1], why the quality of life of affected patients is of increasing relevance. For the last two decades, sentinel lymph node biopsy (SLNB) has been the standard surgical procedure for evaluating axillary status in patients with breast cancer and clinically node-negative (cN0) status [2]. The SLNB procedure causes less postoperative morbidity than axillary lymph node dissection (ALND); however, it is still associated with lymphedema, arm pain/numbness, and quality-of-life reduction [3]. Furthermore, in 70%–80% of cases [4] SLNB will prove negative without cancer cells in the sentinel lymph nodes, and surgical axillary intervention will have no therapeutic benefit.

Multiple recent studies have presented prediction models for noninvasive staging of axillary nodal (N) status with the long-term aim of replacing SLNB for subgroups of patients with breast cancer [5-17]. Only routinely and preoperatively available data should be used for a feasible noninvasive diagnosis of axillary N status aimed at clinical implementation. Most published models include postoperative variables from surgical specimens, including pathological tumor size [10, 14], estrogen receptor (ER) status [5, 7, 13, 16], progesterone receptor (PR) status [5, 7], human epidermal growth factor receptor 2 (HER2) status [5, 7, 10, 16], proliferation index Ki67 value [5, 7, 13], Nottingham histological grade (NHG) [5, 7, 8, 12], histological type [5, 7, 8, 12], and lymphovascular invasion (LVI) [6, 7, 11].

ER, PR, HER2, and Ki67 showed moderate to very good concordance between core needle biopsy (CNB) and surgical specimens [18]. Therefore, these variables have the potential as preoperative predictors of lymph node status. Similarly, NHG and histological type showed more than 70% [19] and 80% [20] concordance rates, respectively, for the same comparison. However, LVI is challenging to evaluate on preoperative CNB because of the limited amount of tissue sample, and a high failure rate of 30% has been reported [21]. Along with tumor size, LVI status is the most important clinicopathological predictor of N status [22]. Although preoperative evaluation of LVI remains a challenge, an accurate preoperative assessment of LVI is needed to predict N status.

Imaging of the breast and axilla can be used to assess preoperative tumor size and extract other features related to N status. Standard imaging modalities in the diagnostic workup of breast cancer are mammography and ultrasound (US) of the breast and axilla; therefore, data from these imaging modalities can be obtained routinely. Several models have been developed using US features [5, 10, 11, 16, 17]. However, the US is operator-dependent; therefore, it is not reproducible, which limits its utility in prediction models. In addition, prediction models using other imaging modalities or combinations, such as US and magnetic resonance imaging (MRI) [9], positron emission tomography combined with US [13], MRI [14], contrast-enhanced spectral mammography (CESM) [15], and US combined with computed tomography [16] lack clinical feasibility.

Nomograms have been developed based on postoperative, non-imaging, and pathological data. Li et al. [8] showed an internal validation area under the receiver operating characteristics curve (AUC) of 0.718 (95% CI: 0.714–0.723) when predicting lymph node metastasis including tumor size, NHG, and histological type. The discriminatory performance of the Memorial Sloan-Kettering Cancer Center nomogram [22] for prediction of sentinel lymph node metastasis, developed based on n=3786 patients, decreased significantly from an AUC of 0.75 in the internal validation to an AUC of 0.67, 95% CI: 0.63–0.72, when externally validated in a Dutch population (n=770) [23]. Furthermore, the Skåne University Hospital (SUS) nomogram [6], a logistic regression model based on 800 patients in Lund, Sweden, aiming to predict negative sentinel lymph nodes, had an internal validation AUC of 0.74 (95% CI: 0.70–0.79). The nomogram was temporally (n=1318) and geographically (n=1621) externally validated with an AUC of 0.75 (95% CI: 0.70–0.81) and an AUC of 0.73 (95% CI: 0.70–0.76), respectively [24].

In 2019, Dihge et al. [7] predicted axillary nodal status in patients with cN0 breast cancer using a multilayer perceptron (MLP) model for noninvasive lymph node staging (NILS) based on 15 clinical and postoperative pathological predictors. The NILS concept includes logistic regression and machine learning models for noninvasive staging of the axilla, aiming at a web interface implementation to be used in clinical practice. Similar to previous nodal prediction models, pathological tumor size and LVI were the top-ranked predictors in the original (MLP) NILS model [7]. Training and internal cross-validation were performed on the same n=800 patients as in Dihge et al. [6] and provided a prediction of the disease-free axilla. In addition, the possible clinical benefit of using the model to detect patients who were least likely to benefit from SLNB was assessed. Surgical axillary lymph node staging could have been avoided in 27% of patients given a false negative rate (FNR) of 10%, corresponding to the accepted FNR for SLNB [25]. Although the benefit of replacing logistic regression with machine learning in clinical prediction models is not given [26], the MLP model outperformed the multivariable logistic regression model, given its discriminatory performance.

This study primarily aimed to externally validate the original NILS model presented in 2019 [7] and develop a new N model in a large population-based cohort of routinely collected data from The Swedish National Quality Registry for Breast Cancer (NKBC). In addition, it secondarily aimed to develop an LVI model and assess how the overall predictive performance of the N model is affected by applying the LVI model for missing values. To the best of our knowledge, this is the first LVI model to be incorporated into an N model. This study was conducted in concordance with the Transparent Reporting of a multivariate prediction model of Individual Prognosis Or Diagnosis to develop and validate prediction models [27].

## METHODS

### Study population

Three datasets with non-overlapping populations were used for model development and evaluation. The inclusion criteria for all three cohorts were female patients with invasive primary breast cancer and cN0 axilla scheduled for primary surgical treatment, with excision of the breast tumor by total mastectomy or partial mastectomy and axillary staging by SLNB. In addition, exclusion criteria for the three cohorts were male, previous ipsilateral breast or axillary surgery, bilateral cancer, previous neoadjuvant therapy, ductal carcinoma *in situ* only, missing pathological-anatomical diagnosis (PAD) tumor size, tumor size > 50 mm (T3), a tumor growing into the chest wall or skin (T4), metastatic disease (stage IV breast cancer), patients with clinically node-positive disease, and missing or incongruent data for axillary surgery and/or lymph node status.

The three datasets originated from different periods. Dataset I (n=995) comprised consecutive patients diagnosed with primary breast cancer at the SUS Lund, Sweden, between January 2009 and December 2012. Data were extracted from medical records and pathology reports, with a final cohort size of n=761 (Figure S1, *Appendix*). For Dataset I, a quality assessment scheme was performed to ensure accurate histopathological reporting and internal quality control of the retrieved data from medical records. Dataset II (n=23 264) was a large population-based cohort of a breast cancer registry for external validation and development of a new N model. It consisted of patients with primary breast cancer from all breast cancer treatment units in Sweden, included in the NKBC register during 2014–2017, with a final cohort size of n=18 633 (Figure 1). Löfgren et al. [28] examined the data quality of NKBC in 2019 and reported high validity and coverage of 99.9% for 2010–2014. Dataset III (n=598) comprised consecutive patients with primary breast cancer surgically treated in Malmö or Helsingborg, Sweden, between 2020 and 2019–2020, respectively. Data were, similar to those of Dataset I, extracted from medical records and pathology reports. The final cohort size was 525 patients (Figure S2, *Appendix*). The data extraction for Cohort III was validated and monitored by an independent researcher according to a specific quality assurance protocol [29]. The sample size calculation for validating the NILS concept has been published previously [29].

**Figure 1.**
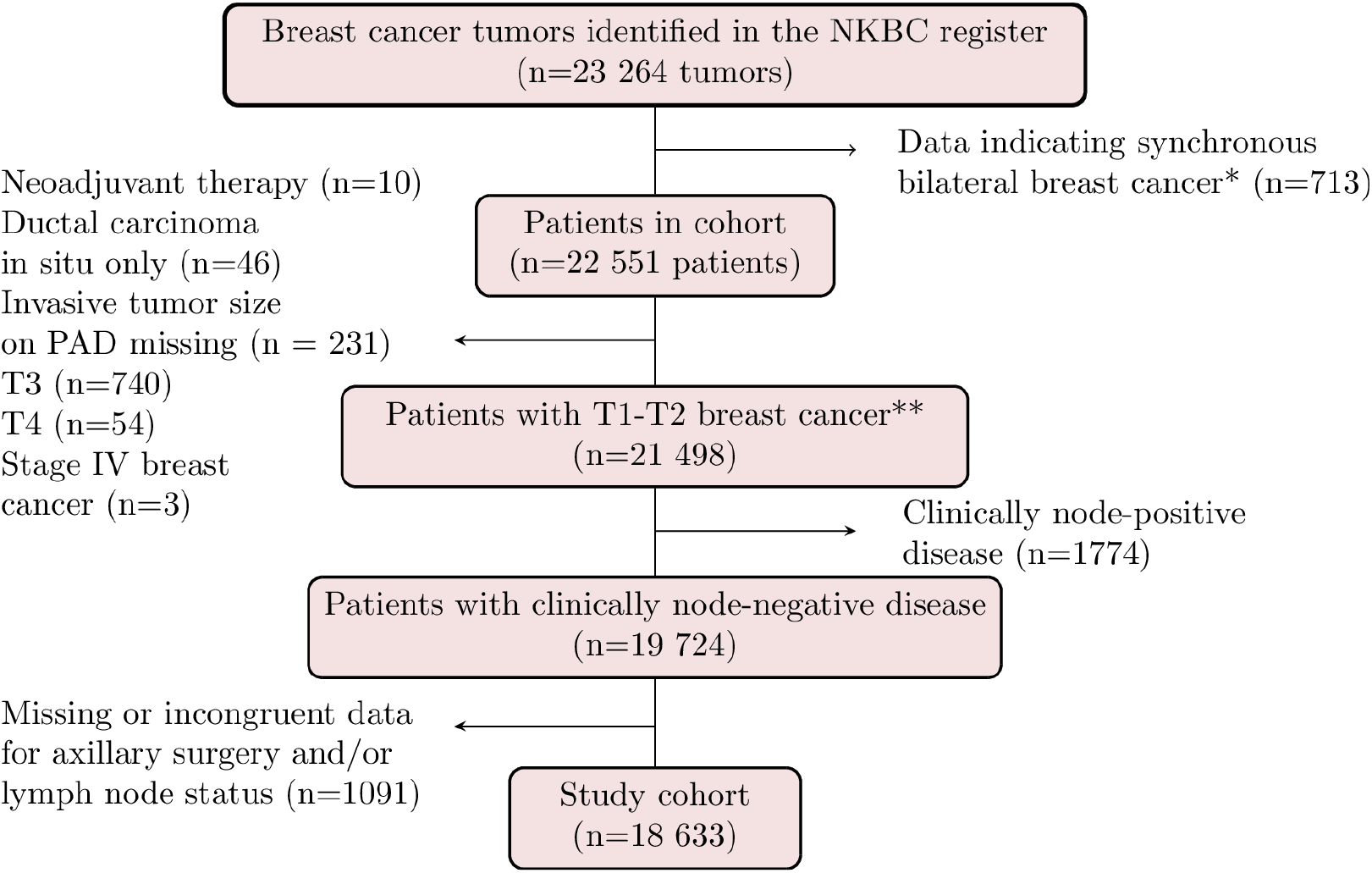
Patient selection for Cohort II. *Including records with the same information on age, mode of detection, hospital, and date of diagnosis, but with different laterality. ** Note that 31 patients were excluded by two of the six criteria in this step. NKBC, Swedish National Quality Register for Breast Cancer; PAD, pathological-anatomical diagnosis.

### Endpoints

The following two endpoints were assessed: pathological N status (node-negative (N0) *vs*. node-positive (N+) disease) and pathological LVI status (LVI-positive *vs*. LVI-negative disease). Lymph node involvement was defined as metastatic infiltration > 0.2 mm in the lymph nodes; therefore, also patients with only nodal micrometastasis were included in the study and categorized as N+. LVI positivity was defined as the presence of tumor cells within endothelium-lined lymphatic or blood-vascular channels [30]. A board-certified specialist in clinical pathology assessed both endpoints on surgical breast specimens according to the national guidelines for pathology [30].

### Data availability and preprocessing

The original NILS model [7] included the following variables available preoperatively: age at diagnosis, body mass index (BMI), tumor laterality, mode of detection (mammographic screening or symptomatic presentation), menopausal status, tumor localization (centrally or 1–12 o’clock position), and variables assessed on surgical breast specimens: largest pathological tumor size, tumor multifocality assessed by pathology, histological type, NHG, LVI status, ER status, PR status, HER2 status, and Ki67 labeling index. Access to variables differed in the large population-based register Cohort II (Table 1) compared with that of Cohort I and III (Table S1-S2, *Appendix*, respectively) with data extracted from medical records. BMI and tumor localization data were not routinely registered in NKBC, and these two variables were excluded. In addition, the inclusion of tumor characteristics and lymph node status in the contralateral breast and axilla violated the assumption of independent samples, and patients with bilateral tumors were excluded. Although the information on LVI status was missing in Cohort II, a separate prediction model for LVI status was developed in Cohort I because of its importance in predicting N status [7, 22]. All variables were defined and preprocessed as described by Dihge et al. [7], except for the histological type. In cohorts I and II, the histological type was categorized into the following three groups: no special type, lobular, and other/mixed. In Cohort III, data on other/mixed histological type was regrouped, and the mixed histological type was set as missing.

**Table 1.** Patient and tumor characteristics for Cohort II. The number of missing values is shown for non-complete case variables.

|  | All patients (n=18 633) | N0 (n=14 829) | N+ (n=3804) |
| --- | --- | --- | --- |
| **Age (years), median (range)** |  |  |  |
|  | 65 (22-95) | 65 (22-95) | 63 (23-94) |
| **Menopausal status** |  |  |  |
| Premenopausal |  |  |  |
|  | 3336 (19%) | 2515 (18%) | 821 (23%) |
| Postmenopausal |  |  |  |
|  | 14 224 (81%) | 11 457 (82%) | 2767 (77%) |
| Missing |  |  |  |
|  | 1073 | 857 | 216 |
| **Mode of detection** |  |  |  |
| Mammographic screening |  |  |  |
|  | 10 816 (58%) | 8992 (61%) | 1824 (48%) |
| Symptomatic presentation |  |  |  |
|  | 7777 (42%) | 5802 (39%) | 1975 (52%) |
| Missing |  |  |  |
|  | 40 | 35 | 5 |
| **Tumor size (mm), median (range)** |  |  |  |
|  | 15 (1-50) | 14 (1-50) | 19 (1-50) |
| **Multifocality** |  |  |  |
| Absent |  |  |  |
|  | 15 537 (84%) | 12 730 (86%) | 2807 (74%) |
| Present |  |  |  |
|  | 3061 (16%) | 2075 (14%) | 986 (26%) |
| Missing |  |  |  |
|  | 35 | 24 | 11 |
| **Histological type** |  |  |  |
| No special type (NST) |  |  |  |
|  | 14 322 (77%) | 11 325 (76%) | 2997 (79%) |
| Lobular |  |  |  |
|  | 2387 (13%) | 1862 (13%) | 525 (14%) |
| Other invasive,<br>including mixed<br>types |  |  |  |
|  | 1924 (10%) | 1642 (11%) | 282 (7%) |
| **NHG** |  |  |  |
| I |  |  |  |
|  | 4112 (23%) | 3600 (25%) | 512 (14%) |
| II |  |  |  |
|  | 9672 (54%) | 7595 (53%) | 2077 (56%) |
| III |  |  |  |
|  | 4643 (26%) | 3458 (24%) | 1185 (32%) |
| Missing |  |  |  |
|  | 206 | 176 | 30 |
| **ER status** |  |  |  |
| Negative (< 1%) |  |  |  |
|  | 1490 (8%) | 1199 (8%) | 291 (8%) |
| Positive (≥ 1%) | | | |
|  | 16 423 (92%) | 13 053 (92%) | 3370 (92%) |
| Missing |  |  |  |
|  | 720 | 577 | 143 |
| **PR status** |  |  |  |
| Negative (< 1%) |  |  |  |
|  | 2672 (15%) | 2182 (16%) | 490 (14%) |
| Positive (≥ 1%) | | | |
|  | 14 973 (85%) | 11 839 (84%) | 3134 (86%) |
| Missing |  |  |  |
|  | 988 | 808 | 180 |
| **HER2 status** |  |  |  |
| Negative |  |  |  |
|  | 16 288 (89%) | 12 989 (89%) | 3299 (88%) |
| Positive |  |  |  |
|  | 2082 (11%) | 1627 (11%) | 455 (12%) |
| Missing |  |  |  |
|  | 263 | 213 | 50 |
| **Ki67 (%)**, median<br>(range) |  |  |  |
|  | 20 (0-100) | 20 (0-100) | 24 (1-100) |
| Missing |  |  |  |
|  | 133 | 123 | 10 |

### Study design

This was an observational diagnostic study. Because of the absence of information on LVI status in Cohort II, three N models trained in Cohort I (N-LVI_present^I^, N-LVI_imputed^I^, and N-LVI_absent^I^, Figure 2) were developed to externally validate the original NILS model [7]. Each of the three models had different access to values for LVI status. When applicable, missing data on the LVI status were imputed using an LVI model (LVI model in Figure 2). The model N-LVI_present^I^ was developed using only patients with documented LVI status (n=613 patients in Cohort I). For the model N-LVI_imputed^I^, patients with missing values for LVI status (n=148 patients) had these predicted using the LVI model, and the model was trained on all n=761 patients in cohort I. The model N-LVI_absent^I^ was developed without access to LVI status in all 761 patients in Cohort I. The LVI model was developed based on 613 patients in Cohort I with documented LVI status.

**Figure 2.**
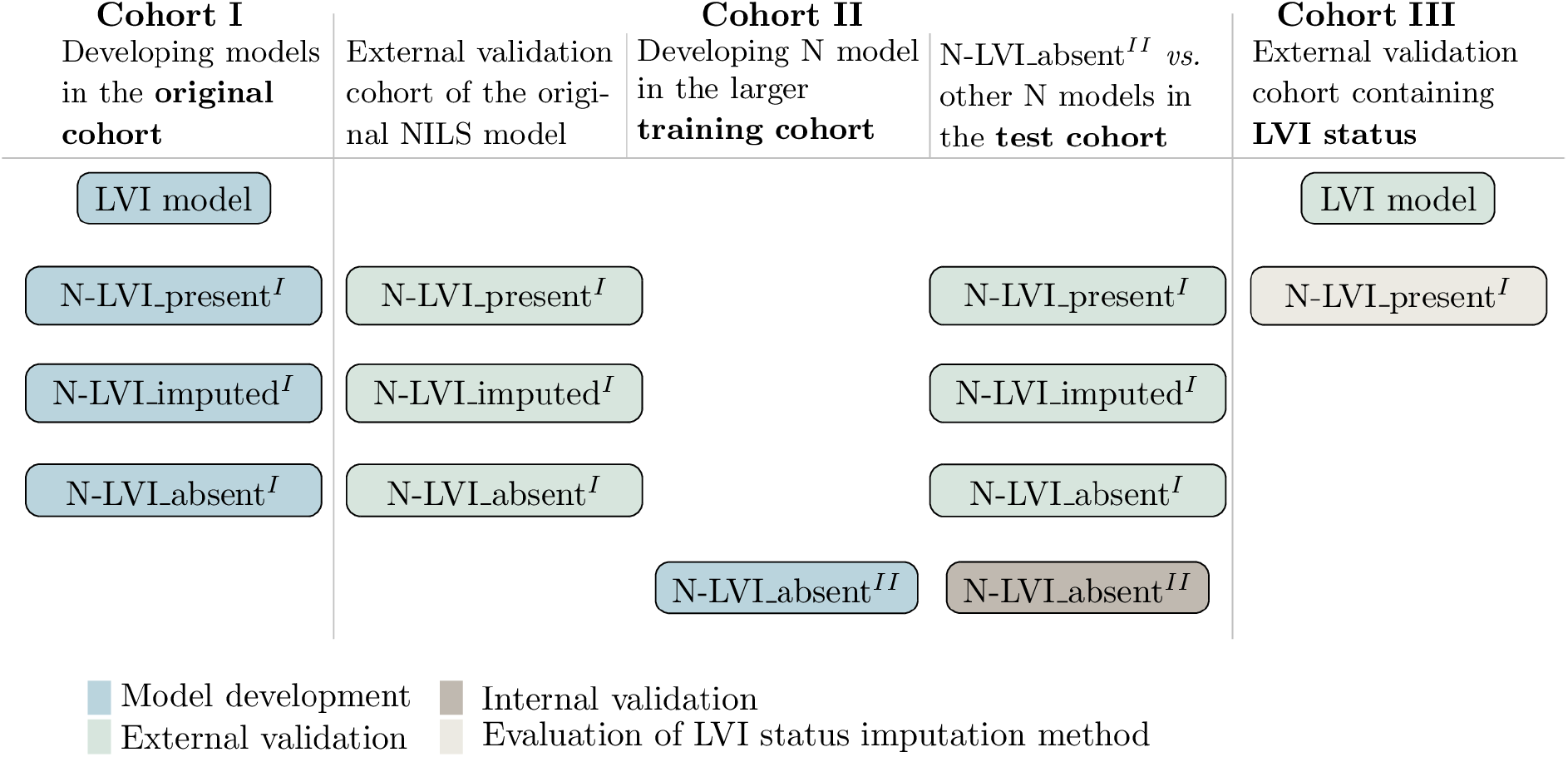
Models developed and evaluated in the study. Three N models were developed to account for the lack of data on LVI status in Cohort II. The external validation was made in Cohort II (NKBC) (n=18 633). A new N model was developed in the training cohort (n=14 906) of Cohort II, and its performance was compared to that of the three other N models in the test cohort (n=3727) of Cohort II. An LVI model was developed to predict LVI status for patients without documented LVI status in Cohort I, and the LVI model was externally validated in Cohort III. In addition, different alternatives for LVI imputation were tested in the model N-LVI_present^I^ in Cohort III. LVI, lymphovascular invasion; N, nodal; NKBC, Swedish National Quality Register for Breast Cancer.

The three available cohorts enabled us to externally validate the original NILS model [7] and investigate the effect of imputed values of LVI status on N model predictions. Imputations by the LVI model were further evaluated in the model N-LVI_present^I^ (see subheading *LVI status model evaluation*). The considerably larger size of Cohort II also enabled the development of a new N model (N-LVI_absent^II^; Figure 2) in a large population-based cohort.

Cohort II was categorized into a training and a test dataset of 80%/20% stratified by N status to compare the performance of the model N-LVI_absent^II^ with that of N models N-LVI_present^I^, N-LVI_imputed^I^, and N-LVI_absent^I^. The model N-LVI_absent^II^ was developed using the training dataset (n=14 906), whereas the test dataset (n=3727) was set aside for comparison with the other developed N models.

### Model development and selection

The process of training the LVI model and the four N models was similar to that in Dihge et al. [7] but with minor modifications owing to different access to data, as presented above. Briefly, an ensemble MLP was developed for each examined hyperparameter combination, and every network in the ensemble was trained using 5-fold cross-validation, stratified by the endpoint distribution. The mean validation AUC of each ensemble was compared to identify the hyperparameter combination that yielded the highest validation AUC. One difference from the original model development was using random search instead of grid search, where each learning algorithm was assigned randomly selected hyperparameters, given a range of values. This hyperparameter optimization method is more efficient than iterating over all possible hyperparameter combinations [31].

### Missing data

The three cohorts had between 1%–2% missing values and 72%–90% complete-case patients (Tables S3–S5, *Appendix*). Missing LVI status were assumed to be missing at random conditional on the other predictors, and other values were assumed to be missing completely at random. In the original NILS model, missing data were handled using multiple random imputation. In this study, missing data were imputed either by multiple random imputation or by the LVI model.

All cases with missing LVI status values were predicted using the LVI model. During the development of the model N-LVI_imputed^I^, the LVI model was used to predict the LVI status of 148 patients lacking information on LVI status in Cohort I at the beginning of each fold in the 5-fold cross-validation. For each training epoch, the LVI status was set to LVI positive or negative, given the probability of the prediction. Missing values among other variables were imputed using multiple random imputation where a missing value was randomly replaced by a value in the present data distribution for the corresponding variable. The procedure was repeated at the beginning of each training epoch.

### LVI model evaluation

To evaluate the LVI model developed in Cohort I, three types of imputations of LVI status were compared with the original values for LVI status in Cohort III. The comparison was made using the N status predicted by the N-LVI_present^I^ model. Subsequently, the three types of imputation were 1) the probability predicted using the LVI model, 2) the corresponding category (LVI-positive/LVI-negative) given the probability of the prediction, and 3) the corresponding category of the prediction given a cut-off of 0.3, matching the distribution of the LVI predictions in the internal cross-validation to that of the development cohort.

The imputation option yielding the highest validation AUC for N status, calculated as the mean of the N-LVI_present^I^ model’s predictions over 25 imputed datasets, was chosen for the imputation of LVI status in Cohort II. Calibration curves of the observed versus mean predicted probabilities were used to visualize the LVI model calibration.

### N model evaluation

The N model validation AUC was calculated as the mean of the AUCs over 25 datasets imputed for missing values and the LVI status was imputed by the LVI prediction model for each dataset when applicable. In addition, a secondary outcome for the N models was the proportion of patients that could be omitted from SLNB while maintaining the FNR at 10% (the generally accepted FNR of SLNB [25]). Set beforehand, developing an N model identifying candidate patients for SLNB omission in every fifth patient with cN0 breast cancer was to be considered successful.

Model predictions were recalibrated to the prevalence in the external validation cohort to account for the different N status distributions of Cohorts I and II [32]. In addition, calibration curves of the observed *vs*. mean predicted probabilities were used to visualize the model calibration. Finally, decision curves [33] were analyzed to examine the standardized clinical benefit [34] of the N models, where the threshold probabilities were set to the range of the acceptable level for the FNR (0%–10%).

### Software and hardware

All parts of the study were conducted using Python 3.9.7 (Python Software Foundation) [35] and TensorFlow 2.6.0 [36], with an Intel® Core™ i7-8700K CPU @ 3.70GHz or a GPU (2xGeForce RTX 2080) computer.

### Ethics

The Regional Ethics Committee at Lund University, Sweden, approved Cohort I for the study (LU 2013/340). Cohorts II and III were approved for the study by the Swedish Ethical Review Authority (2019–02139) and (2021-00174), respectively.

## RESULTS

### LVI model to predict missing values of LVI status for NILS

The LVI model was trained on 613 patients in Cohort I and evaluated in Cohort III (n=525) (Figure 2 and Table S5, *Appendix*). The model had an internal cross-validation AUC of 0.799 (95% CI: 0.751–0.846) and an external validation AUC of 0.747 (95% CI: 0.694–0.799) (Figure 3). In addition, the LVI model showed good calibration in external validation (Figure S3, *Appendix*). The final architecture for the LVI and N models can be found in the Appendix (Table S7).

**Figure 3.**
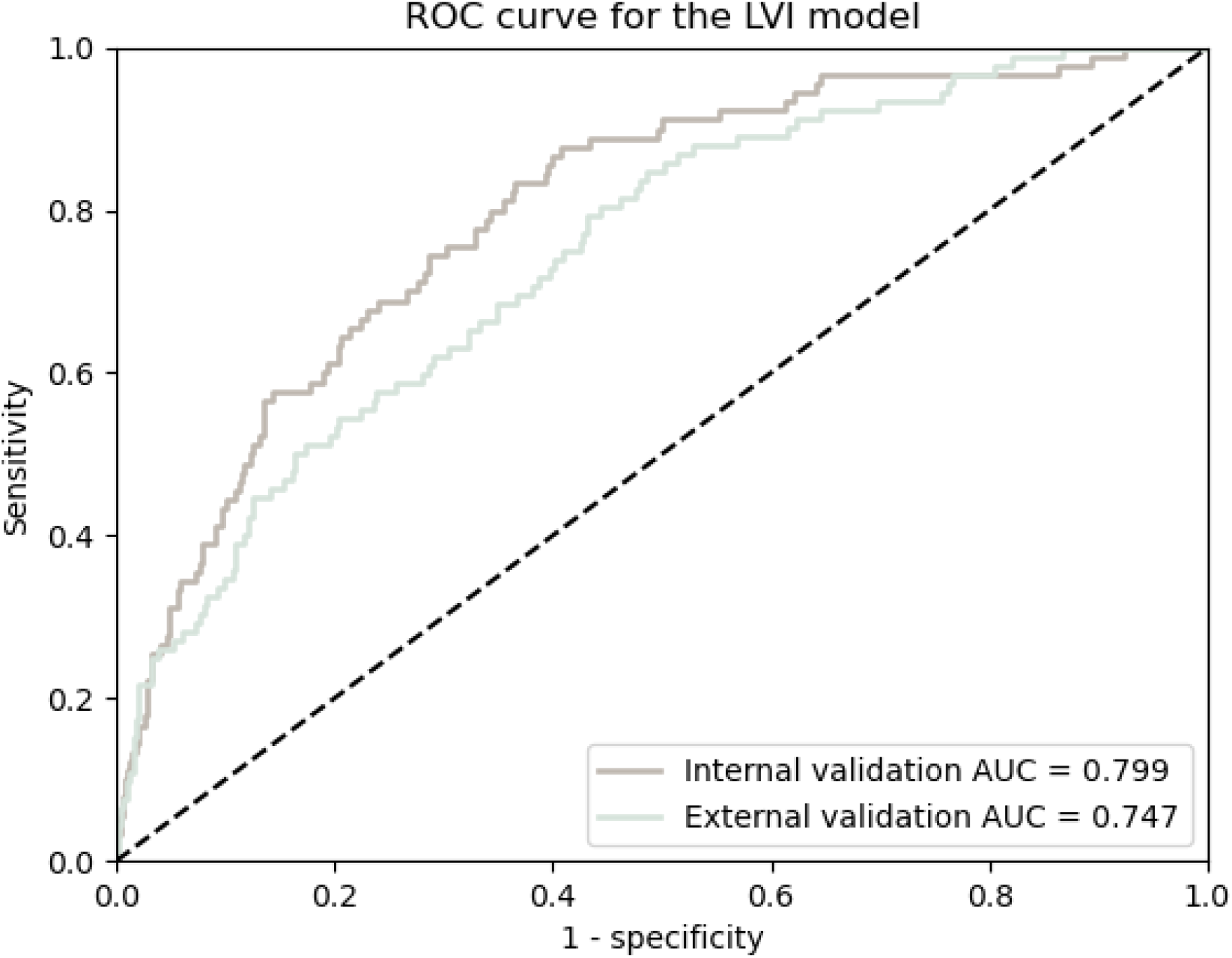
The LVI model had a discriminatory performance AUC of 0.799 (95% CI 0.751–0.864) in the internal validation and an AUC of 0.747 (95% CI 0.694–0.799) in the external validation. ROC, receiver operating characteristic; AUC, area under the ROC curve.

All alternatives for LVI imputation were evaluated in Cohort III using the N model N-LVI_present^I^. The model N-LVI_present^I^ imputed with probabilistically drawn categorical values of LVI status performed slightly better than the other options (Table 2); therefore, this type of LVI imputation was subsequently used.

**Table 2.** AUC for the N status predictions of the model N-LVI_present^I^ for different strategies for imputing values of LVI status. The highest AUC, except when using the original LVI values, was obtained when imputing LVI status using the probabilistic imputation, the reason this method was chosen for LVI imputation in the rest of the analysis. AUC, area under the receiver operating characteristic curve; LVI, lymphovascular invasion; N, nodal.

|  | Original<br>LVI status | LVI status<br>imputed by the<br>predicted<br>probability | LVI status<br>imputed by<br>probabilistically<br>categorical<br>imputation | LVI status<br>imputed by<br>categorical<br>imputation with<br>threshold 0.3 |
| --- | --- | --- | --- | --- |
| **N-**<br>**LVI_present<sup>I</sup>,**<br>**AUC (95%**<br>**CI)** |  |  |  |  |
|  | 0.750<br>(0.704-0.795) | 0.737<br>(0.689 – 0.783) | 0.740<br>(0.693-0.784) | 0.738<br>(0.691-0.783) |

### External validation of the original NILS model

To externally validate the original NILS model in Cohort II without information on LVI status, three N models (N-LVI_present^I^, N-LVI_imputed^I^, and N-LVI_absent^I^) were developed for Cohort I (n=761), as shown in Figure 2. The original NILS model was internally cross-validated with an AUC of 0.740 (95% CI: 0.723–0.758) [7]. In the present external validation in Cohort II (n=18 633), both the N-LVIpresent^I^ and N-LVIimputed^I^ models reached an AUC of 0.686 (95% CI: 0.677–0.695) (Figure 4, *left*). Furthermore, upon validation, the model N-LVIabsent ^I^ reached an AUC of 0.699 (95% CI 0.690–0.708). The classification performance of all N models is summarized in the Appendix (Table S8).

**Figure 4.**
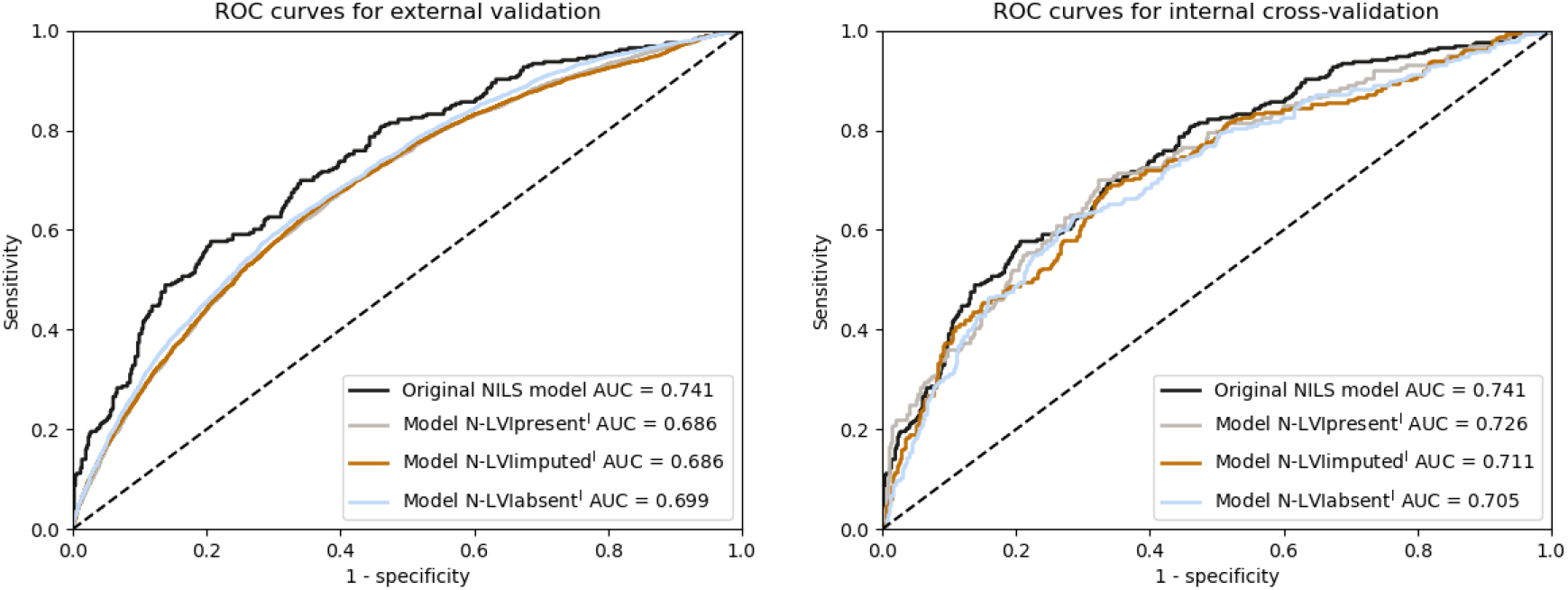
The ROC curve for the external validation (left) and the internal validation (right) of the original NILS model in Dihge et al. [7]. The models of this study had access to slightly different variables and a different number of patients in the training cohort than that of the original NILS model. The models N-LVI_present^I^ and N-LVI_imputed^I^ both included LVI status, whereas N-LVI_absent^I^ did not. Note that the original model was cross-validated with AUC 0.740 in [7], which was an average of five runs. The ROC curve of the original NILS model is in this figure represented by the run closest to the mean; AUC 0.741. LVI, lymphovascular invasion; ROC, receiver operating characteristic; AUC, area under the ROC curve; NILS, noninvasive lymph node staging.

### The impact of the LVI model on the overall N status predictions

The internal validation of the N models showed a higher performance for models N-LVI_present^I^ and N-LVI_imputed^I^ using LVI status (AUC 0.726, 95% CI: 0.681–0.768 and AUC 0.711, 95% CI: 0.762–0.750, respectively), compared to that of model N-LVI_absent^I^ not including LVI status (AUC 0.705, 95% CI 0.665–0.744) (Figure 4, *right*). For external evaluation of the models N-LVI_present^I^ and N-LVI_imputed^I^, the LVI model was used to predict the LVI status in Cohort II.

When externally validated in Cohort II (n=18 633), the models N-LVI_present^I^, N-LVI_imputed^I^, and N-LVI_absent^I^ showed similar performances (Figure 4, *left*). Therefore, the rest of the external validation focused on the model developed without access to LVI status, N-LVI_absent^I^. In the calibration plot, the model N-LVI_absent^I^ demonstrated slightly lower predictions than the true values in the external validation (Figure S4, *Appendix*).

However, when transforming the predictions in relation to the prevalence of N0 in the validation cohort, the calibration of the model N-LVI_absent^I^ was satisfactory.

### Comparison between developed N models

The fourth N status model, N-LVI_absent^II^, was developed in NKBC, a large population-based cohort. The cohort was considerably larger (training cohort n=14 906) than the development cohort for the other three N models and the original NILS model [7] (Cohort I). The test cohort of Cohort II (n=3727), set aside before the development of model N-LVI_absent^II^, was used to compare the performance of the developed N models. The models N-LVI_present^I^, N-LVI_imputed^I^, and N-LVI_absent^I^ reached AUC of 0.684 (95% CI: 0.663–0.705), 0.685 (95% CI: 0.663–0.706), and 0.696 (95% CI: 0.676–0.717), respectively (Figure 5). The model N-LVI_absent^II^ reached a slightly higher AUC of 0.709 (95% CI: 0.688–0.729). The calibration plot for the model N-LVI_absent^II^ is shown in the Appendix (Figure S5).

**Figure 5.**
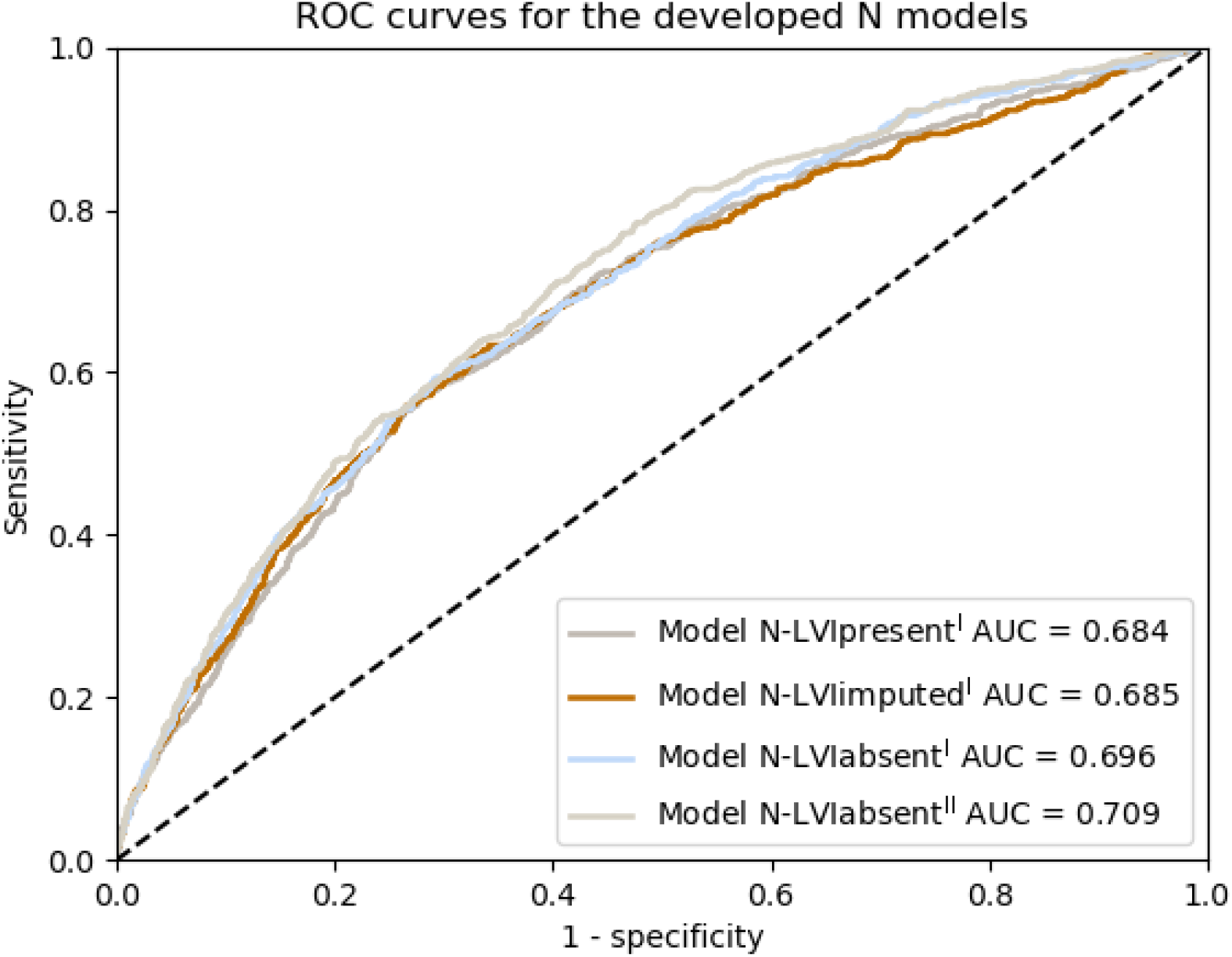
Validation in the test cohort (n=3727) of Cohort II for the N models N-LVI_present^I^, N-LVI_imputed^I^, N-LVI_absent^I^, and the new N model N-LVI_absent^II^ developed in a larger cohort. N, nodal; ROC, receiver operating characteristic; AUC, area under the ROC curve.

### Potential clinical utility

External validation of the N models before recalibration showed potential in sparing approximately 20% of cN0 patients from axillary surgery when using an FNR of < 10%. When recalibrating the predictions for the model N-LVI_absent^I^, the number decreased to approximately 16%. However, the new N model N-LVI_absent^II^ developed in Cohort II could potentially spare 24% of patients with cN0 breast cancer from SLNB. The standardized decision curve analyses (Figure 6) specifically showed the range of predictions where patients could benefit from utilizing the two prediction models. The standardized decision curve analysis for the original predictions of N-LVI_absent^I^ before recalibration is shown in the Appendix (Figure S5).

**Figure 6.**
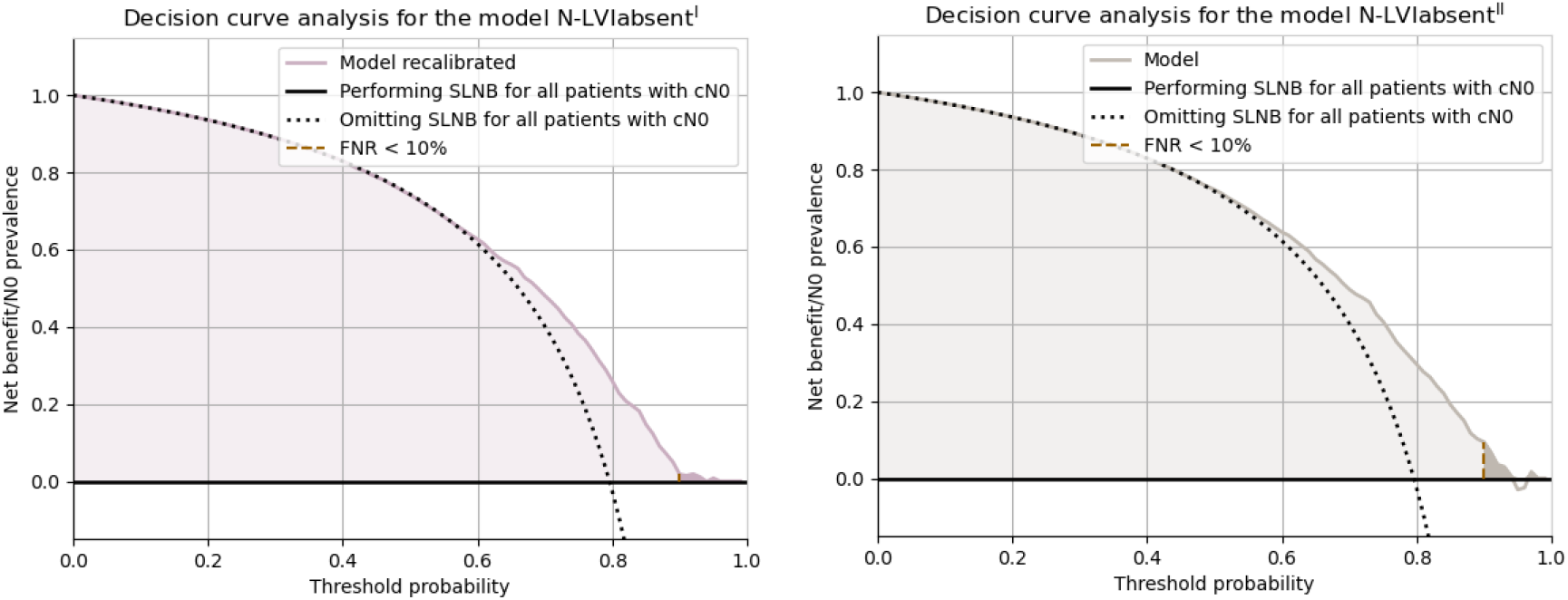
Decision curves showing the standardized net benefit of the model N-LVI_absent^I^ (recalibrated) (left) and the model N-LVI_absent^II^ (right). The black horizontal line represents the scenario of all patients being diagnosed as node-negative; hence, no SLNB is performed. The colored function represents the diagnosis by the model. The golden, vertical (dashed) line at a threshold of approximately 0.9, separating the lighter color from the darker, shows the threshold for FNR < 10%. When all patients are considered node-positive and diagnosed through SLNB, the standardized net benefit is, by definition, zero. Notably, the darker, colored area does not represent the patients spared from surgery. Rather, it displays the standardized net benefit of the model where FNR < 10%. cN0, clinically node negative; FNR, false negative rate; SLNB, sentinel lymph node biopsy.

## DISCUSSION

### Principal results

The proportion of patients diagnosed with early-stage breast cancer is increasing [4]. Along with improvements in adjuvant therapy, surgical treatment is becoming more conservative. Most patients with early-stage breast cancer have benign lymph nodes and would benefit from preoperative noninvasive staging of the axilla [3, 4]. In this study, we externally validated a previously published N model [7] in a national, large population-based register cohort (n=18 633) without access to LVI status and developed a new N model within the same cohort. Notably, the discriminatory performance (AUC 0.709, 95% CI: 0.688–0.729) of the new N model (N-LVI_absent^II^) developed in the large population-based cohort demonstrated that routine clinicopathological register data can be used to develop an N model to identify 24% of patients with cN0 for whom surgical axillary staging could be circumvented. The model developed in Cohort I without access to data on LVI status (N-LVI_absent^I^) achieved an AUC of 0.699 (95% CI: 0.690–0.708) and the potential to omit 16% of patients from SLNB. The use of fewer variables and, in some cases, fewer patients was expected to result in a slight decrease in the performance of the models in this study compared with that of the original model.

### Comparison with prior studies

Multiple studies have investigated the discriminatory performance of nomograms for predicting N status using retrospective clinicopathological data alone or in combination with imaging features [6, 8-17]. We aimed to externally validate and further develop a diagnostic tool for the noninvasive staging of N status using only routinely available clinicopathological data to improve the clinical utility of the model. Li et al. [8] and Gao et al. [12] developed nomograms using solely clinicopathological data that can be obtained preoperatively.

However, the studies did not specify whether the data were extracted from the preoperative or postoperative setting. Li et al. had the advantage of a very large cohort (n=184 532); unfortunately, combining external validation data with parts of the development cohort resulted in inaccurate external validation (AUC 0.718). Gao et al. developed a nomogram based on 6314 patients with external validation on 503 patients, where the shift from training and internal validation to external validation increased from an AUC of 0.715 and 0.688 to an AUC of 0.876, respectively. This large discriminatory increase in external validation is unexpected and warrants questioning the model’s validity.

One possibility for the transition from postoperative to preoperative variables is the use of imaging features. Mao et al. [15] developed a nomogram using CESM reported lymph node status and a radiomics signature to predict axillary lymph node status. In addition, the nomogram was externally validated on only n=62 patients with an AUC of 0.79 (95% CI: 0.63–0.94). Using only features that can be obtained preoperatively is an advantage in Mao et al.’s study [15]. However, additional larger external validation is needed to confirm the results of the study. Furthermore, CESM is not part of the mammography screening program or routine workup for suspicious breast malignancies, limiting the study’s clinical feasibility. Bove et al. [5] developed a support vector machine (SVM) classifier for clinical data and one SVM for radiomics data to predict N status. They used soft voting, which implies combining the probabilities of each prediction in the two models. They chose the prediction with the highest total probability, which resulted in an AUC of 0.886 on the hold-out test set.

Combining pre- and postoperative variables is a limitation of the study, and the axillary US is an operator-dependent imaging modality. However, the results show the potential for using imaging features in machine learning models for noninvasive staging of N status. The SVM classifier had an AUC of 0.739 using only postoperative clinicopathological data, similar to that of the original NILS model [7]. However, both the training (n=114) and test (n=28) datasets were small; therefore, a larger external validation is needed to confirm the results.

In this study, the LVI model, trained using only routine clinicopathological variables and developed to increase the feasibility of the NILS models in the preoperative setting, had an external validation AUC of 0.747 (95% CI: 0.694–0.799). To the best of our knowledge, this is the first LVI model to be incorporated into an N model. Preoperative assessment of LVI on CNB is challenging, and several models have been developed to predict the LVI status. For example, Shen et al. [37] developed a logistic regression model for the LVI status using clinicopathological variables (n=392). Although the model reached an AUC of 0.670 (95% CI: 0.607–0.734) in the training dataset, it was not further validated. In addition, others have investigated the importance of radiomics features for predicting LVI status, for example, digital mammography features [38] with LVI prediction specificity of 98.8% in the development cohort and MRI features [39] with an AUC of 0.732 in the test dataset.

However, while highlighting the potential for predicting LVI status using radiomics, the data used are not part of the diagnostic workup for breast cancer, limiting clinical feasibility.

Despite the AUC of 0.747 for the LVI model in this study, the imputation of values for LVI status did not improve the discriminatory performance of the N models in the large population-based register cohort (NKBC). This may indicate that the uncertainty in the LVI model’s predictions was still too large for the LVI model to be of benefit when predicting N status. Nevertheless, the reliability and/or distribution of data, such as multifocality, may change in the preoperative setting [40], which could change the prerequisites for predicting LVI status. Given the growing evidence on the significance of LVI status as a predictor of axillary N status [7, 11, 22, 41], further evaluation of the presented LVI model is warranted.

### Potential clinical utility

Omitting SLNB in subgroups of patients is consistent with the American Society of Clinical Oncology guidelines from 2021 [2], stating that SLNB is optional for all patients ≥ 70 years old with cN0, ER+, and HER2-if the patient received adjuvant endocrine therapy. In this study, using only routine clinicopathological data, the models developed without access to LVI status in Cohort I (recalibrated) and Cohort II presented the potential to spare 16%–24% of patients with cN0 from SLNB, irrespective of age and tumor subtype. In addition, a health-economic study concluded that the NILS model is cost-effective [42]. If lymphedema is considered to negatively impact the patients’ quality of life, the NILS model also showed a net health gain [42].

### Strengths and limitations

Criticism has been raised against the use of small sample sizes in the development and external validation of machine learning models in oncology, as well as the poor handling of missing data [39]. Accordingly, we aimed to externally validate the original NILS model [7] in a nationwide and large population-based register cohort (n=18 633) and to develop a new NILS model within this larger cohort (n=14 906). Using a large population-based register cohort is advantageous in the following two ways: 1) its consecutive nature constitutes a good approximation of the true distributions of the population, and 2) it demonstrates the reality of data handling where input data will comprise missing values and occasional mistakes in documentation. Importantly, we have shown that register data can be used to develop an N model with equally satisfactory results as when using more curated data, including the LVI status. Our external validation of the original NILS model [7] was performed in a temporally, geographically, and domain-wise different cohort from the original development cohort. We presented calibration and net benefit curves to demonstrate the utility of the models. In addition, the 1091 patients in Cohort II with missing or incongruent data for axillary surgery and/or lymph node status (Figure 1) showed a similar distribution of clinical variables (data not shown) as the final study population of Cohort II. Therefore, there was no indication of selection bias.

Another strength of our study is the thorough management of missing data using both the LVI model and multiple random imputation. Our comprehensive handling of missing values may increase the utility of N models in a clinical preoperative setting. It also showed that for the discriminatory performance in N staging, the manner the predictions of LVI status were presented to an N model was of minor importance. However, this requires further investigation in the preoperative setting and/or utilization of an LVI model with even higher discriminatory performance to completely rule out the potential advantage of MLP LVI predictions in NILS.

However, this study had some limitations. First, the models were developed using a combination of variables available before and after surgery to externally validate the original NILS model [7] which is based on preoperative and postoperative variables. Further development of the NILS concept is an ongoing validation of the NILS model, using exclusively preoperative variables [29]. Second, the generalizability of the LVI and N models developed in Cohort I can be affected by the smaller size of the development cohorts which can be considered a weakness of the study. Therefore, regularization of the networks and 5-fold cross-validation were used to minimize overfitting. The drop in performance from the internal to external validation was small for all models, which is a clear strength of our findings.

Recalibration was performed for the model developed without access to LVI status in cohort I (N-LVI_absent^I^) because of the different prevalence of benign lymph nodes in Cohorts I and II (65% pN0 *vs*. 80% pN0). No recalibration was performed for the LVI model because the prevalence of a positive LVI status was similar in Cohorts I and III. Notably, when transforming the N status predictions in relation to the new prevalence, the calibration and the overall net benefit of the model N-LVI_absent^I^ improved, while the fraction of patients to be spared from SLNB decreased. Therefore, to potentially increase the number of candidate patients to be omitted from SLNB, an important future development of the model could be to evaluate it using partial AUC [43] or concordant partial AUC [44]. The model selection is then based on the model’s performance under specific conditions, for example, FNR < 10%, which could optimize the model performance for patients most likely to benefit from the prediction. Another option is to investigate the modification of the loss function when training the MLP to optimize the algorithm for the largest number of patients to be omitted from SLNB while keeping the FNR < 10%.

An additional strength of this study was the use of three disjoint cohorts for model development and validation. After model development, two patients in Cohort I were incorrectly classified as N0 instead of N+. However, these two patients corresponded to less than 1% of the cohort and did not affect the overall results. Cohort II demonstrated high validity and a high coverage of key variables [28]. An independent researcher validated and monitored Cohort III according to a specific quality assurance protocol to ensure well-characterized data. All variables, except one, were defined in coherence; the mixed histological type was categorized as missing in Cohort III. However, this should have a limited effect on the results since mixed histological type is rare (approximately 5%) [45]. We excluded bilateral patients not to risk dependency and information leakage between the two tumors and/or nodal statuses. The exclusion limits the target group to a minor extent, as bilateral cancers are generally diagnosed in less than 5% of patients [46].

### Future studies

Future steps include a prospective external validation of the NILS concept in a larger cohort and an evaluation of the incorporation of LVI predictions in a NILS model in the preoperative setting. External validation of the LVI model in a Norwegian breast cancer cohort is also planned. The feasibility of using register data for prediction modeling demonstrates the possibility of using larger and less-curated databases in machine learning models for NILS.

Implementing neural network models that are equal or superior to linear models allows extending the model to more complex data that cannot be handled by logistic regression in end-to-end learning. This enables less human interference, simpler implementation, and models to optimize the entire task. Therefore, to potentially improve the discriminatory performance of noninvasive staging of lymph nodes for future clinical implementation, additional types of data conferring to the knowledge of lymphatic spread should ideally be investigated. Imaging features are both preoperatively available and have shown high discriminatory performance in nodal prediction models [5, 9-11, 13-15]. In addition, molecular subtypes are associated with the outcome as well as N status and the difficult-to-treat triple-negative subtype has the lowest risk of nodal metastasis compared to luminal tumors [6]. Consequently, models based on gene expression analysis have shown potential in correctly identifying N0 patients in specific subtypes of breast cancer, such as luminal-A [47], ER+/HER2- [48] and triple-negative tumors [49], to capture additional aspects of lymphatic spread, such as immune signatures. Gene expression data have also shown the potential to increase the number of candidate patients to be omitted from SLNB when combined with clinicopathological data compared to predicting N status using clinicopathological data alone [50]. Therefore, planned extensions of the NILS model include mammography images and gene expression data, mainly focusing on molecular subtypes and immune signatures.

## Conclusions

We externally validated the original NILS model [7] in a large population-based register cohort, with a discriminatory performance of 0.699 (95% CI: 0.690–0.708). Prediction of LVI status did not improve the performance of the N model, despite its documented importance in prediction of axillary stage. A new MLP model for predicting N status was developed in a large population-based register cohort, demonstrating the feasibility of developing a prediction model for noninvasive nodal staging using register data comprising only variables available in the preoperative setting and, notably, no information on LVI status (AUC 0.709, 95% CI: 0.688–0.729). Therefore, future studies include evaluating the LVI model in the preoperative setting, the ongoing preoperative validation of the NILS concept, and extend the NILS model with preoperative and routinely available data such as mammography images and gene expression data.

## Supporting information

TRIPOD checklist

## Data Availability

The raw datasets are available upon reasonable request due to restrictions such as privacy and ethical restrictions.

## Data availability

The raw datasets are available upon reasonable request due to restrictions such as privacy and ethical restrictions. The data are not publicly available due to these restrictions. The code is available upon reasonable request.

## Abbreviations

ALND: axillary lymph node dissection
AUC: area under the receiver operating characteristic curve
BMI: body mass index
CESM: contrast-enhanced spectral mammography
CNB: core needle biopsy
cN0: clinically node-negative
ER: estrogen receptor
FNR: false negative rate
LVI: lymphovascular invasion
MLP: multilayer perceptron
MRI: magnetic resonance imaging
HER2: human epidermal growth factor receptor 2
N: nodal
NHG: Nottingham histological grade
NILS: noninvasive lymph node staging
NKBC: Swedish National Quality Register for Breast Cancer
PAD: pathological-anatomical diagnosis
PR: progesterone receptor
ROC: receiver operating characteristic
SLNB: sentinel lymph node biopsy
SUS: Skåne University Hospital
SVM: support vector machine
US: ultrasound

