## Supplemental figures and tables for "Noninvasive Staging of Lymph Node Status (NILS) in Breast Cancer using Machine Learning: External Validation and Further Model Development"

### Appendix

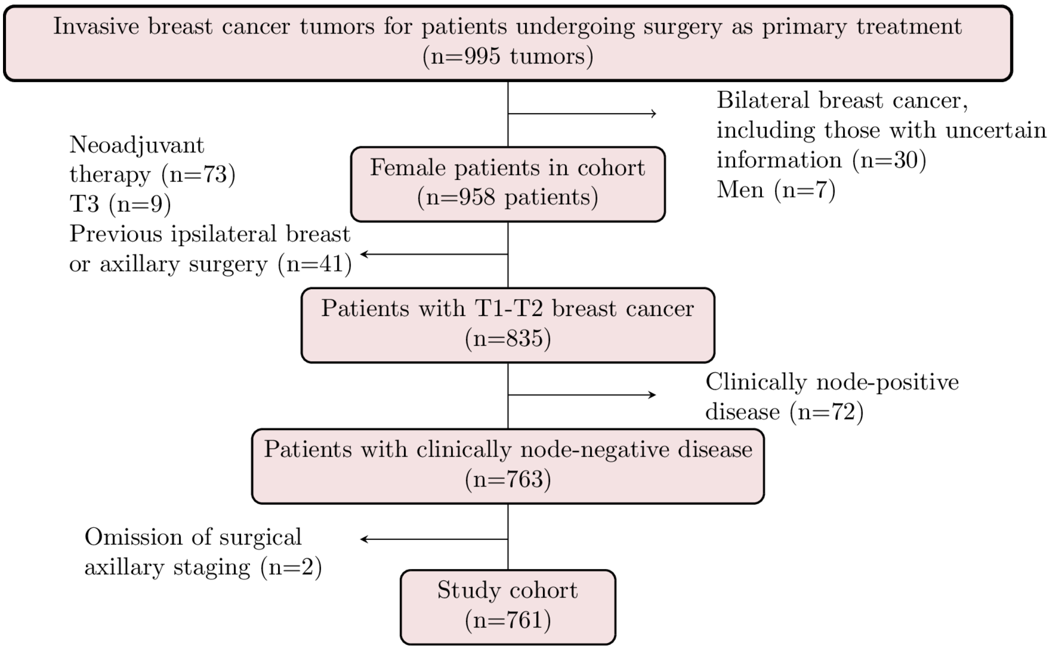

*Figure S1. Patient selection for Cohort I.*

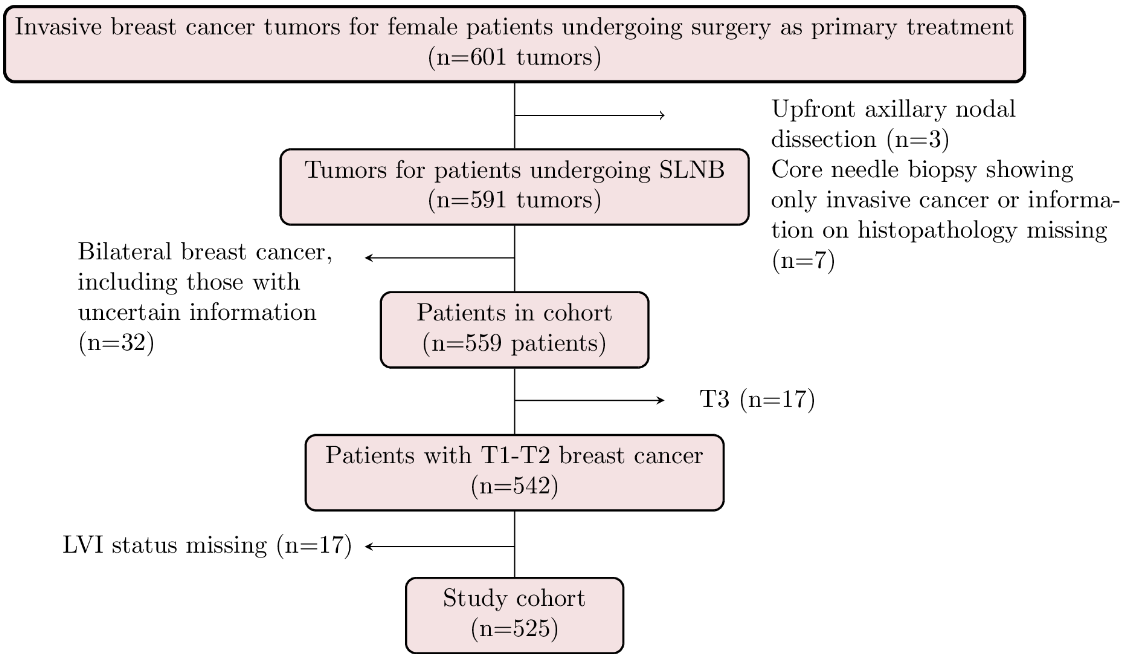

*Figure S2. Patient selection for Cohort III. LVI, lymphovascular invasion, SLNB; sentinel lymph node biopsy.*

*Table S1. Patient and tumor characteristics in Cohort I. The number of missing values is shown for non-complete case variables.*

| | All patients (n=761) | N0 (n=497) | N+ (n=264) |
|---|---|---|---|
| **Age** (years), median (range) | | | |
| | 65 (24-92) | 66 (33-91) | 64 (24-92) |
| **Menopausal status** | | | |
| Premenopausal | | | |
| | 128 (18%) | 70 (15%) | 58 (23%) |
| Postmenopausal | | | |
| | 591 (82%) | 397 (85%) | 194 (77%) |
| Missing | | | |
| | 42 | 30 | 12 |
| **Mode of detection** | | | |
| Mammographic screening | | | |
| | 447 (59%) | 315 (63%) | 132 (50%) |
| Symptomatic presentation | | | |
| | 314 (41%) | 182 (37%) | 132 (50%) |
| **Tumor size** (mm), median (range) | | | |
| | 15 (0.5-50) | 13 (0.5-50) | 18 (0.9-50) |
| Missing | | | |
| | 1 | 0 | 1 |
| **Multifocality** | | | |
| Absent | | | |
| | 575 (76%) | 402 (81%) | 173 (66%) |
| Present | | | |
| | 186 (24 %) | 95 (19%) | 91 (34%) |
| **Histological type** | | | |

| | | | |
|---|---|---|---|
| No special type (NST) | | | |
| | 613 (81%) | 397 (80%) | 216 (82%) |
| Lobular | | | |
| | 93 (12%) | 57 (11%) | 36 (14%) |
| Other invasive, including mixed types | | | |
| | 55 (7%) | 43 (9%) | 12 (5%) |
| **NHG** | | | |
| I | | | |
| | 186 (25%) | 136 (29%) | 50 (19%) |
| II | | | |
| | 346 (47%) | 224 (47%) | 122 (47%) |
| III | | | |
| | 221 (30%) | 130 (27%) | 91 (35%) |
| Missing | | | |
| | 8 | 7 | 1 |
| **ER status** | | | |
| Negative (< 1%) | | | |
| | 67 (9%) | 52 (11%) | 15 (6%) |
| Positive (≥ 1%) | | | |
| | 692 (91%) | 443 (89%) | 249 (94%) |
| Missing | | | |
| | 2 | 2 | 0 |
| **PR status** | | | |
| Negative (< 1%) | | | |
| | 119 (16%) | 87 (18%) | 32 (12%) |
| Positive (≥ 1%) | | | |
| | 640 (84%) | 408 (82%) | 232 (88%) |
| Missing | | | |
| | 2 | 2 | 0 |

| **HER2 status** | | | |
|---|---|---|---|
| Negative | | | |
| | 618 (88%) | (89%) | 209 (86%) |
| Positive | | | |
| | 83 (12%) | (11%) | 33 (14%) |
| Missing | | | |
| | 60 | 38 | 22 |
| **Ki67** (%), median (range) | | | |
| | 15 (0-92) | 14 (0-92) | 17 (1-81) |
| Missing | | | |
| | 42 | 28 | 14 |
| **Lymphovascular invasion** | | | |
| Absent | | | |
| | 523 (85%) | 386 (93%) | 137 (69%) |
| Present | | | |
| | 90 (15%) | 27 (7%) | 63 (32%) |
| Missing | | | |
| | 148 | 84 | 64 |

*Table S2. Patient and tumor characteristics in Cohort III. The number of missing values is shown for non-complete case variables.*

| | All patients (n=525) | N0 (n=401) | N+ (n=124) |
|---|---|---|---|
| **Age** (years), median (range) | | | |
| | 66 (29-91) | 66 (29-89) | 65 (34-91) |
| **Menopausal status** | | | |
| Premenopausal | | | |

| | | | |
|---|---|---|---|
| | 70 (14%) | 51 (14%) | 19 (17%) |
| Postmenopausal | | | |
| | 415 (86%) | 321 (86%) | 94 (83%) |
| Missing | | | |
| | 40 | 29 | 11 |
| **Mode of detection** | | | |
| Mammographic screening | | | |
| | 318 (61%) | 252 (63%) | 66 (53%) |
| Symptomatic presentation | | | |
| | 207 (39%) | 149 (37%) | 58 (47%) |
| **Tumor size** (mm), median (range) | | | |
| | 14 (1-50) | 13 (4-50) | 20 (1-50) |
| Missing (%) | | | |
| | 9 | 8 | 1 |
| **Multifocality** | | | |
| Absent | | | |
| | 388 (75%) | 310 (78%) | 78 (63%) |
| Present | | | |
| | 130 (25%) | 85 (22%) | 45 (37%) |
| Missing | | | |
| | 7 | 6 | 1 |
| **Histological type** | | | |
| No special type (NST) | | | |
| | 389 (76%) | 291 (74%) | 98 (83%) |
| Lobular | | | |
| | 91 (18%) | 72 (18%) | 19 (16%) |
| Other | | | |
| | 41 (8%) | 36 (9%) | 5 (4%) |

| | | | |
|---|---|---|---|
| Missing | | | |
| | 4 | 2 | 2 |
| **NHG** | | | |
| I | | | |
| | 133 (26%) | 116 (29%) | 17 (14%) |
| II | | | |
| | 294 (57%) | 216 (55%) | 78 (64%) |
| III | | | |
| | 95 (18%) | 67 (17%) | 28 (23%) |
| Missing | | | |
| | 3 | 2 | 1 |
| **ER status** | | | |
| Negative (< 1%) | | | |
| | 34 (6%) | 26 (6%) | 8 (6%) |
| Positive (≥ 1%) | | | |
| | 491 (94%) | 375 (94%) | 116 (94%) |
| **PR status** | | | |
| Negative (< 1%) | | | |
| | 79 (15%) | 63 (16%) | 16 (13%) |
| Positive (≥ 1%) | | | |
| | 444 (85%) | 336 (84%) | 108 (87%) |
| Missing | | | |
| | 2 | 2 | 0 |
| **HER2 status** | | | |
| Negative | | | |
| | 496 (95%) | 383 (96%) | 113 (91%) |
| Positive | | | |
| | 28 (5%) | 17 (4%) | 11 (9%) |
| Missing | | | |
| | 1 | 1 | 0 |
| **Ki67** (%), median (range) | | | |

| | | | |
|---|---|---|---|
| | 21 (2-93) | 20 (2-93) | 29 (5-92) |
| Missing | | | |
| | 1 | 0 | 1 |
| **Lymphovascular invasion** | | | |
| Absent | | | |
| | 433 (82%) | 353 (88%) | 80 (65%) |
| Present | | | |
| | 92 (18%) | 48 (12%) | 44 (35%) |

*Table S3. Data characteristics in development cohorts for nodal status models. No values were missing for the target variable nodal status.*

| | N-LVI_present[Ia] | N-LVI_imputed[Ib] | N-LVI_absent[Ic] | N-LVI_absent[IIc] |
|---|---|---|---|---|
| **Dataset (N)** | | | | |
| | Cohort I (613) | Cohort I (761) | Cohort I (761) | Training cohort II (14 906) |
| **Missing values** | | | | |
| | 67 (1%) | 157 (2%) | 157 (1%) | 2814 (2%) |
| **Complete cases** | | | | |
| | 550 (90%) | 550 (72%) | 626 (82%) | 12 919 (87%) |
| **N0/N+** | | | | |
| | 413/200 (67%/33%) | 497/264 (65%/35%) | 497/264 (65%/35%) | 11 863/3043 (80%/20%) |

[a] Trained on LVI status complete case data.

[b] Trained on data with imputations of missing values of LVI status.

[c] Trained without access to data on LVI status.

*Table S4. Data characteristics in the validation cohorts for the nodal status models. No values were missing for the target variable nodal status.*

| | Cohort II (n=18 633) | Test cohort II (n=3727) |
|---|---|---|
| **Models** | | |
| | N-LVI_present[I]<br>N-LVI_imputed[I]<br>N-LVI_absent[I] | N-LVI_present[I]<br>N-LVI_imputed[I]<br>N-LVI_absent[I]<br>N-LVI_absent[II] |
| **Missing values** | | |
| | 3458 (2%) | 644 (1%) |
| **Complete cases** | | |
| | 16 207 (87%) | 3288 (88%) |
| **N0/N+** | | |
| | 14 829/3804 (80%/20%) | 2966/761 (80%/20%) |

*Table S5. Data characteristics in the development and the evaluation cohort, respectively, for the LVI status model. No values were missing for the target variable LVI. LVI, lymphovascular invasion.*

| | LVI development | LVI evaluation |
|---|---|---|
| **Dataset (N)** | | |
| | Cohort I (613) | Cohort III (525) |
| **Missing values** | | |
| | 67 (1%) | 67 (1%) |
| **Complete cases** | | |
| | 550 (90%) | 459 (87%) |
| **LVI positive/LVI negative** | | |
| | 90/523 (15%/85%) | 92/433 (18%/82%) |

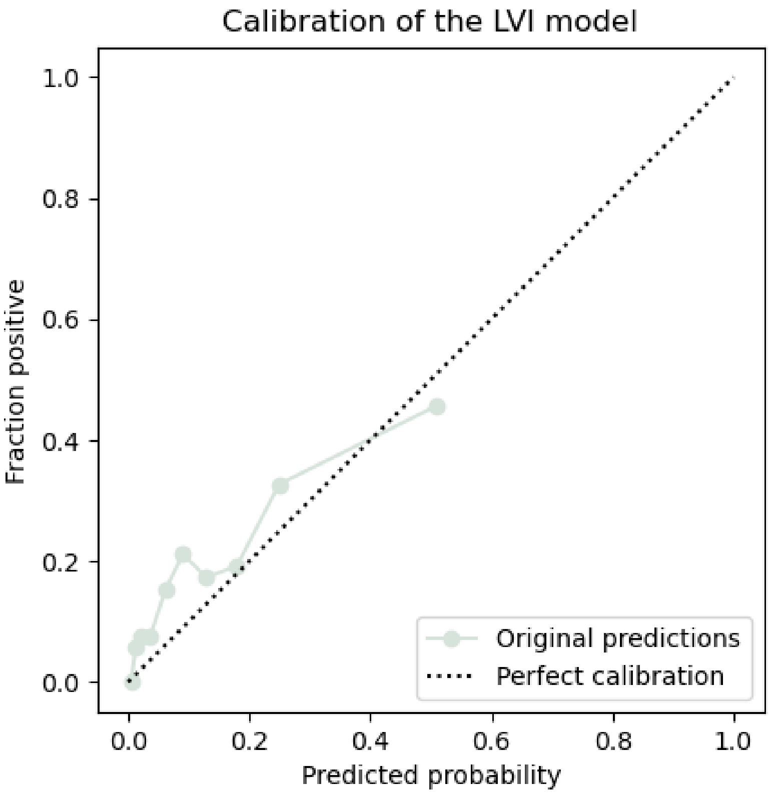

*Figure S3. Calibration of the LVI model in Cohort III.*

*Table S7. Model architecture. All models comprised an input layer of various sizes, one hidden layer, and a one-node output layer. The models were trained during 400 epochs. The loss function was binary cross-entropy.*

| | Learning rate | Number of hidden nodes | L2 regularization | Dropout |
|---|---|---|---|---|
| **N-LVI_present[I]** | | | | |
| | 0.05 | 15 | None | 0.4 |
| **N-LVI_imputed[I]** | | | | |
| | 0.01 | 6 | L2(0.001) | 0 |
| **N-LVI_absent[I]** | | | | |
| | 0.04 | 10 | None | 0.4 |
| **N-LVI_absent[II]** | | | | |
| | 0.009 | 8 | None | 0.5 |
| **LVI model** | | | | |
| | 0.15 | 7 | None | 0.1 |

*Table S8. Sensitivity, specificity, and false negative rate for the N models. The models N-LVI_present[I], N-LVI_imputed[I], and N-LVI_absent[I] were evaluated in Cohort II (n=18 633), and the model N-LVI_absent[II] was evaluated in the test cohort (n=3 727) of Cohort II. FNR, false negative rate.*

| | N-LVI_present[I] | N-LVI_imputed[I] | N-LVI_absent[I] | N-LVI_absent[I] (recalibrated) | N-LVI_absent[II] |
|---|---|---|---|---|---|
| **Sensitivity** | | | | | |
| | 91% | 90% | 90% | 92% | 91% |
| **Specificity** | | | | | |
| | 25% | 27% | 24% | 20% | 30% |
| **FNR** | | | | | |
| | 9% | 9% | 10% | 8% | 9% |

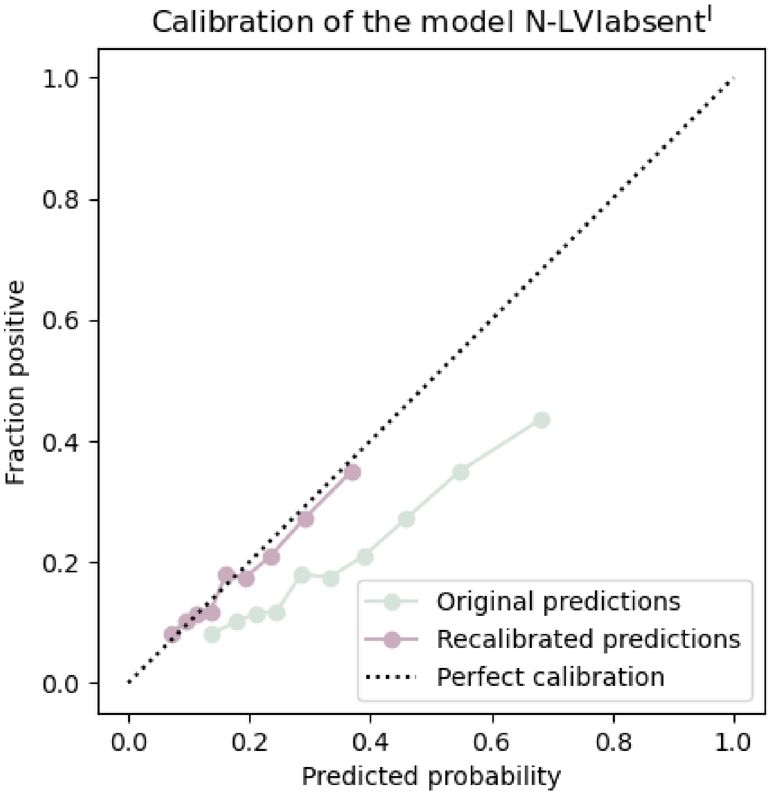

*Figure S4. Calibration and recalibration of the N status model N-LVI_absent[I] in the test cohort of Cohort II. N, nodal.*

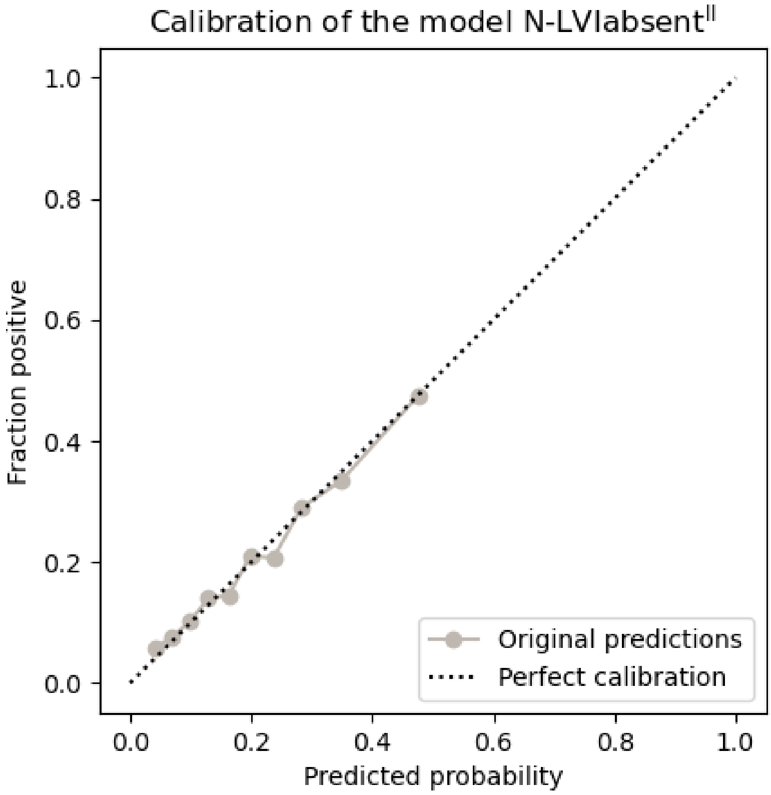

*Figure S5. Calibration of the N status model N-LVI_absent$^{II}$ in the test cohort of Cohort II. N, nodal.*

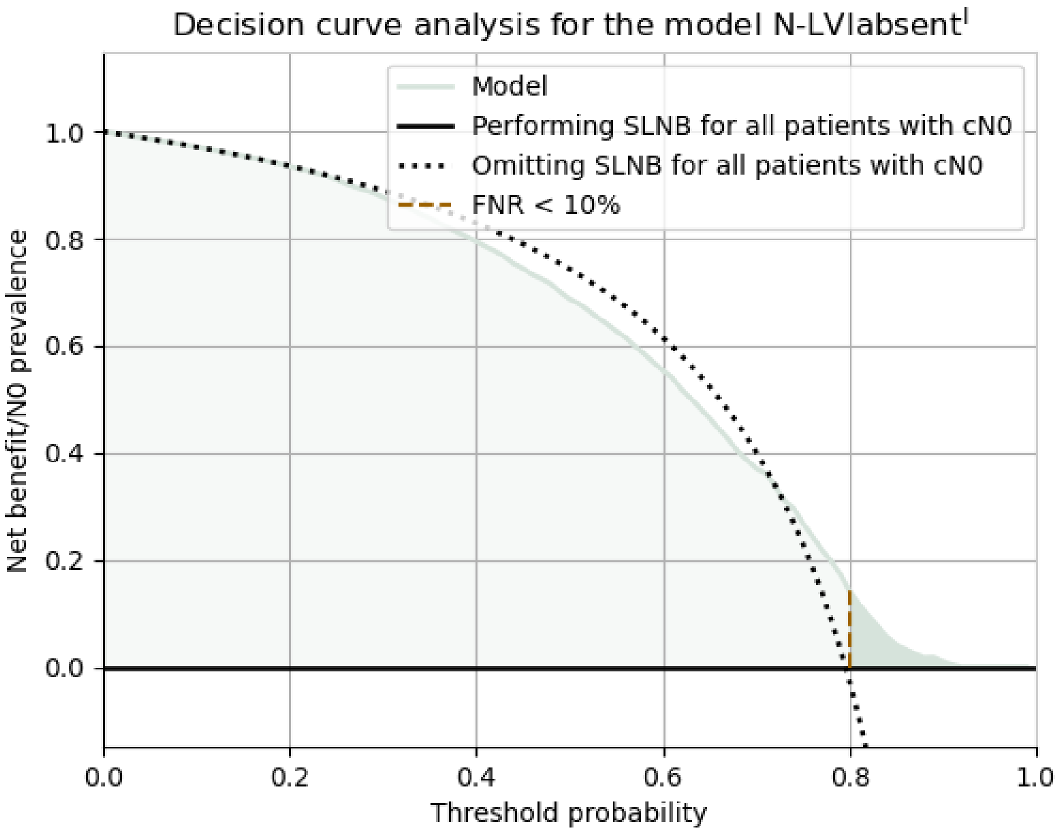

*Figure S6. Standardized decision curve analysis for the original predictions of model N-LVI_absent[I]. The black horizontal line represents the scenario of all patients being diagnosed as node-negative; hence, no SLNB is performed. The colored function represents the diagnosis by the model. The golden, dashed, vertical line separating the lighter color from the darker shows the threshold for FNR < 10%. When all patients are considered node-positive and diagnosed through SLNB, the standardized net benefit is, by definition, zero. Note that the darker, colored area does not represent the patients spared from surgery. Rather, it displays the standardized net benefit of the model where FNR < 10%. cN0, clinically node negative; FNR, false negative rate; SLNB, sentinel lymph node biopsy.*
